# Associations of Race/Ethnicity and Other Demographic and Socioeconomic Factors with Vaccine Initiation and Intention During the COVID-19 Pandemic in the United States

**DOI:** 10.1101/2021.02.16.21251769

**Authors:** Daniel Kim

**Affiliations:** Bouvé College of Health Sciences & School of Public Policy and Urban Affairs, Northeastern University, 360 Huntington Avenue, 413 International Village, Boston MA 02115

## Abstract

**Background:** To date, there has been limited data available to understand the associations between race/ethnicity and socioeconomic and related characteristics with novel coronavirus disease (COVID-19) vaccine initiation and planned vaccination in the United States.

**Methods:** To better characterize COVID-19 vaccinations nationally, we leveraged large cross-sectional surveys conducted between January and March 2021 with relatively complete race/ethnicity and socioeconomic data and nationally-representative of U.S. households to estimate trends in levels of COVID-19 vaccine initiation and vaccine intention. We further used survey data from January and March 2021 in adults aged 18-85 years to analyze the associations between race/ethnicity, education, pre-pandemic household income, and financial hardship during the pandemic and the adjusted odds of: 1) receipt of ≥1 dose of a COVID-19 vaccine; and 2) among those unvaccinated, the definite intention to receive a vaccine, controlling for other demographic and socioeconomic factors.

**Results:** We observed persistent disparities in vaccine initiation for non-Hispanic Blacks, Hispanics, and non-Hispanic multiracial persons, and in vaccine intention for Blacks and multiracial persons, compared to non-Hispanic Whites and non-Hispanic Asians. In late March 2021, the prevalence estimates of Hispanics and Blacks receiving a vaccine were 12 percentage points and 8 percentage points lower than for Whites, respectively. Moreover, both education and income levels exhibited positive dose-response relationships with vaccine initiation (P for trend≤01 and <.001, respectively). Substantial financial hardship was linked to 35-44% lower odds of vaccination (P<.001). The most common reasons for vaccine hesitancy were concerns about side effects and safety, with evidence of higher levels of concerns about vaccine safety among Blacks vs. Whites.

**Conclusions:** In this large, nationally-representative study with relatively complete race/ethnicity and socioeconomic data, we find that being Black non-Hispanic and having the least education and income were each independently associated with a markedly lower likelihood of definitely planning to get vaccinated or having been vaccinated. In the ensuing months of the pandemic, addressing the persevering racial/ethnic and socioeconomic inequities in vaccination due to differential access and vaccine hesitancy is essential to mitigate the pandemic’s higher risks of infection and adverse health outcomes in Hispanic, Black, and socioeconomically-disadvantaged communities.

## Introduction

A recent CDC analysis examined demographic characteristics of nearly 13 million persons who received ≥1 dose of one of two authorized coronavirus disease 2019 (COVID-19) vaccines administered in the United States between December 14, 2020 and January 14, 2021.^1^ Data were collected by vaccination providers and reported to CDC using various reporting methods including immunization information systems. Individuals who were reportedly vaccinated represented approximately 5% of the total U.S. population aged ≥16 years.^1^ Although data on age and sex were reported for more than 97.0% of vaccine recipients, race/ethnicity information was missing for nearly half (49.1%) of individuals, and no socioeconomic data were collected. The study’s investigators and other epidemiologists have called attention to the critical need for more comprehensive reporting of race and ethnicity data to ensure an effective response to racial/ethnic disparities in COVID-19 vaccination, especially among those at disproportionately higher risks for infection and severe adverse health outcomes including racial/ethnic minorities.^1-3^ Furthermore, while the Advisory Committee on Immunization Practices prioritized COVID-19 vaccination of healthcare personnel and long-term care facility residents during the initial phase,^4^ equity considerations need to be factored into vaccine allocation.^5,6^ To better characterize COVID-19 vaccinations nationally, the present study leveraged nationally-representative data with relatively complete race/ethnicity and socioeconomic data to estimate trends in levels of vaccine initiation and intention by race/ethnicity over time, as well as to estimate the adjusted relative odds of vaccine initiation and vaccine intention among adults according to race/ethnicity and other demographic and socioeconomic characteristics.

## Methods

Cross-sectional data were used from non-institutionalized adults aged 18-85 years in the Household Pulse Survey (HPS) by the U.S. Census Bureau, nationally-representative of U.S. households and administered over six periods of the HPS between January 6, 2021 and March 29, 2021, comprising Phase 3 of the HPS.^7^ The online survey response rates ranged from 6.4-7.5%.

The main predictors were race/ethnicity, education, pre-pandemic (2019) household income, and financial hardship. Outcomes consisted of the self-reported: 1) receipt of ≥1 dose of a COVID-19 vaccine; and 2) among those unvaccinated, the intention to definitely receive a vaccine once available to the respondent. Less than 2% of observations in the samples were missing for the items corresponding to these outcomes.

Multivariable logistic regression models were fit to estimate adjusted odds ratios with generalized estimating equations for each outcome to account for survey weights and obtain robust standard errors. Log-Poisson regression was not employed due to bias when estimating relative risks for binary outcomes.^8^ All models included all of the above main predictors, and were also adjusted for age, gender, marital status, household size, children in household, and state of residence. Multiple imputation analysis (using 20 multiply imputed datasets) was employed to handle missing data. All analyses were conducted using SAS (version 9.4; SAS Institute).

## Results

During each HPS period, between 68,348 and 80,567 adults were surveyed, representative of 243 million Americans. Of individuals who reported their COVID-19 vaccination status, race/ethnicity information was missing in 2.1-3.9% of the samples, and was previously handled using hot deck imputation.^7^

*Figure 1* illustrates the trends in racial/ethnic disparities in receiving ≥1 vaccine dose by survey period between January 6, 2021 and March 29, 2021. During the January 6— January 18 period, an estimated 5.9% of Hispanics, 5.9% of non-Hispanic Blacks (Blacks), and 6.2% of non-Hispanic multiracial persons (multiracial persons) had received a vaccine, compared to 8.1% in non-Hispanic Whites (Whites) and 13.3% in non-Hispanic Asians (Asians). The prevalence of vaccine initiation steadily rose over time for all racial/ethnic groups. During the March 17—March 29 period, an estimated 37.8% of Hispanics, 42.4% of Blacks, and 40.1% of multiracial persons had received a vaccine, compared to 50.4% of Whites and 51.7% of Asians.

**Figure 1.**
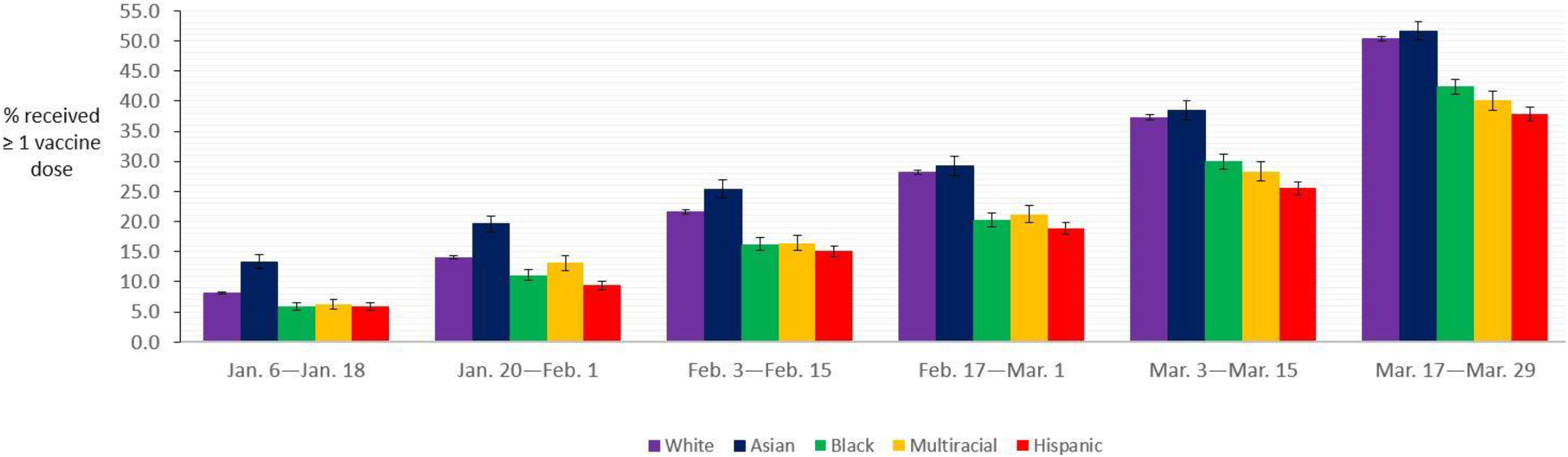
**Estimated Prevalence of Adults Aged 18-85 Years Who Have Received ≥1 Dose of COVID-19 Vaccine by Race/Ethnicity, U.S. Census Bureau Household Pulse Survey, January-March 2021.^a^** Abbreviations: COVID-19, coronavirus disease 2019. ^a^ Aggregate data were drawn from the U.S. Census Bureau Household Pulse Survey public-use data tables for surveys administered between January 6, 2021 and March 29, 2021. All groups except for Hispanics are of non-Hispanic ethnicity. Error bars indicate 95% confidence intervals for prevalence estimates. All estimates are survey weighted.

*Figure 2* displays the time trends in racial/ethnic disparities in the definite intention to get vaccinated among those not yet vaccinated. During the January 6—January 18 period, an estimated 29.6% of Blacks, 47.3% of Hispanics, and 37.3% of multiracial persons had a definite intention to receive a vaccine, compared to 55.5% of Whites and 66.0% of Asians. The prevalence of vaccine intention was higher in subsequent time periods for all racial/ethnic groups except for Whites. During the March 17—March 29 period, an estimated 37.6% of Blacks, 52.9% of Hispanics, and 40.2% of multiracial persons had a definite intention to receive a vaccine, compared to 46.6% of Whites and 71.1% of Asians.

**Figure 2.**
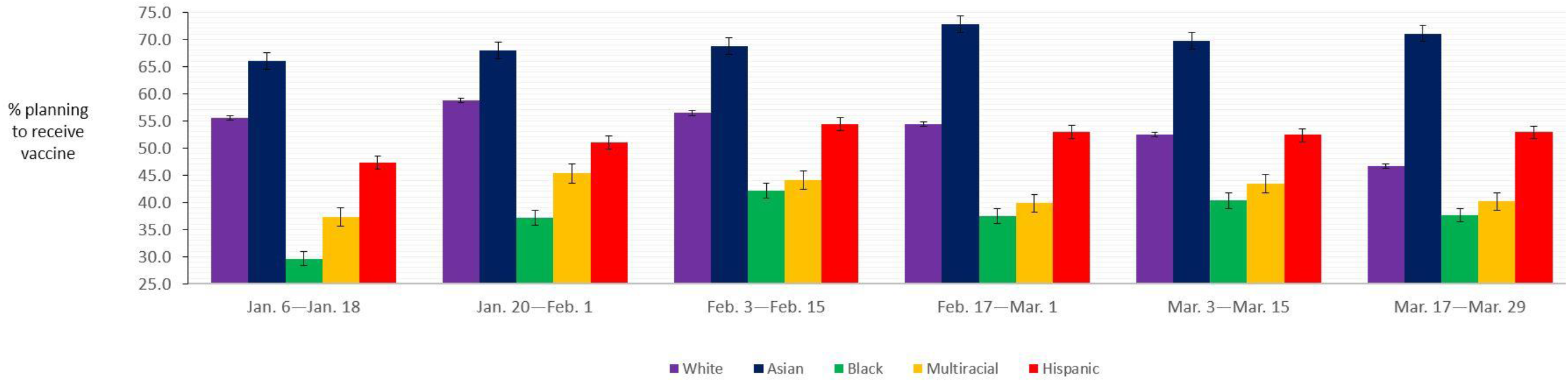
**Estimated Prevalence of Adults Aged 18-85 Years Who Plan to Definitely Get COVID-19 Vaccine by Race/Ethnicity, U.S. Census Bureau Household Pulse Survey, January-March 2021.^a^** Abbreviations: COVID-19, coronavirus disease 2019. ^a^ Aggregate data were drawn from the U.S. Census Bureau Household Pulse Survey public-use data tables for surveys administered between January 6, 2021 and March 29, 2021. All groups except for Hispanics are of non-Hispanic ethnicity. Error bars indicate 95% confidence intervals for prevalence estimates. All estimates are survey weighted.

Based on data from the initial HPS period with vaccination questions (January 6— January 18) and the last HPS period (March 17—March 29), controlling for other demographic and socioeconomic factors, being of Black or multiracial race/ethnicity was not characterized by a higher or lower likelihood of receiving ≥1 vaccine dose than White race/ethnicity (*Table 1*). Meanwhile, Asian race/ethnicity was associated with a higher odds of vaccine initiation than White race/ethnicity in the first and last survey periods (*P*≤.001). Hispanic ethnicity was linked to a greater odds of vaccine initiation than White race/ethnicity in the last survey period (*P*=.001).

**Table 1.**
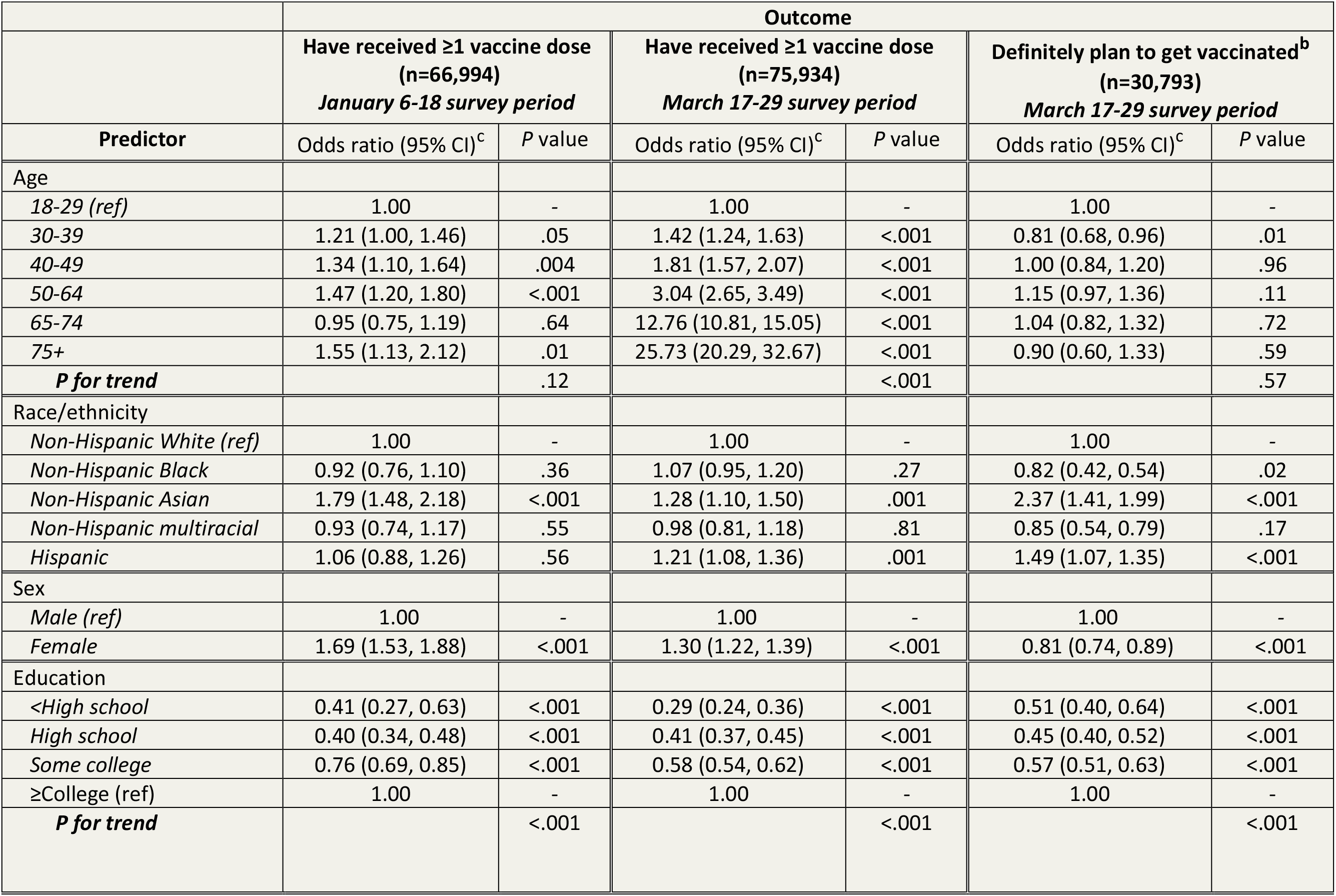
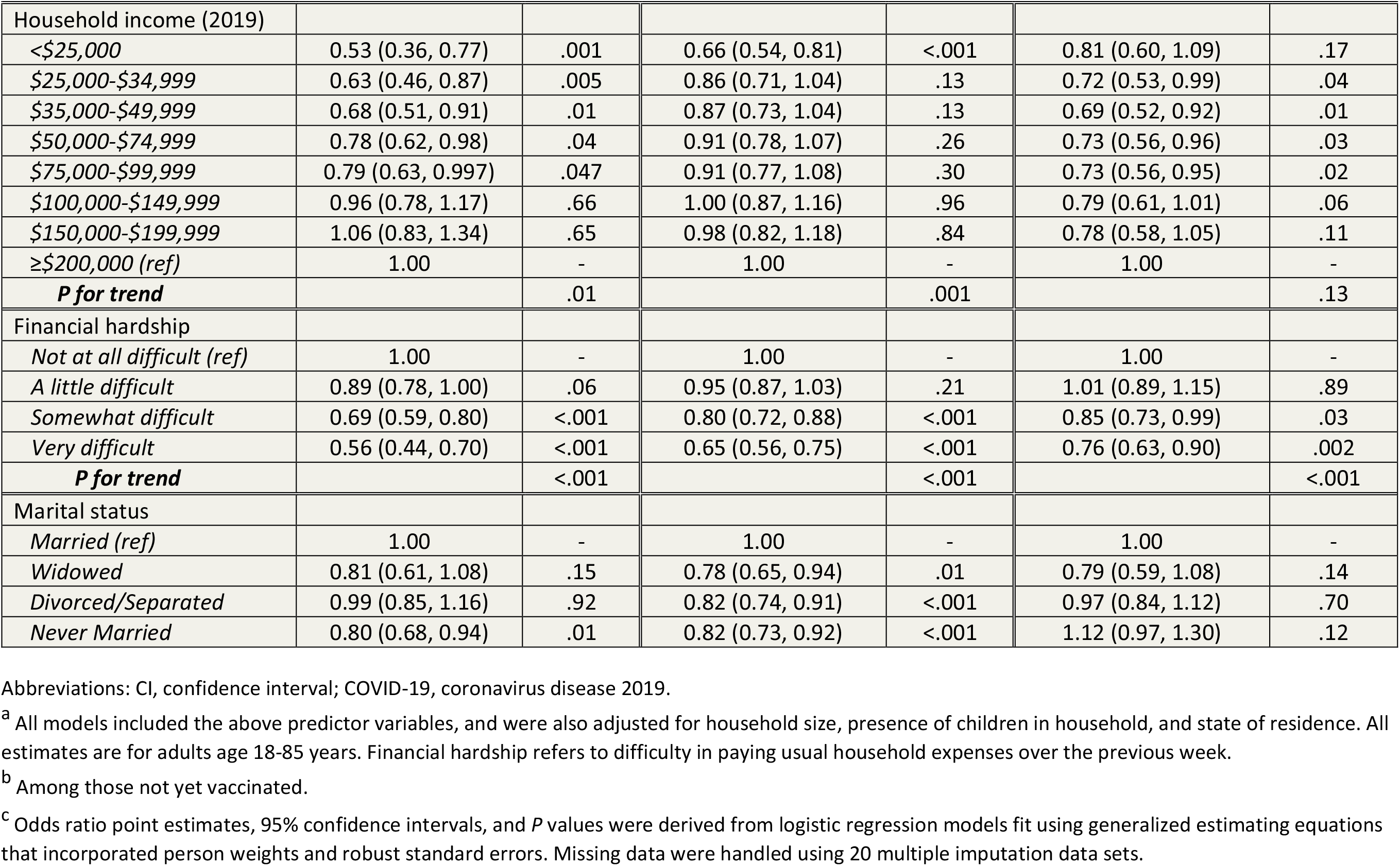
**Demographic and Socioeconomic Predictors of COVID-19 Vaccine Initiation and Vaccine Intention, U.S. Census Bureau Household Pulse Survey, January-March 2021.^a^**

| Predictor | Outcome |  |  |  |  |  |
| --- | --- | --- | --- | --- | --- | --- |
|  | Have received ≥1 vaccine dose<br>(n=66,994)<br>*January 6-18 survey period* |  | Have received ≥1 vaccine dose<br>(n=75,934)<br>*March 17-29 survey period* |  | Definitely plan to get vaccinated<sup>b</sup><br>(n=30,793)<br>*March 17-29 survey period* |  |
|  | Odds ratio (95% CI)<sup>c</sup> | P value | Odds ratio (95% CI)<sup>c</sup> | P value | Odds ratio (95% CI)<sup>c</sup> | P value |
| Age |  |  |  |  |  |  |
| 18-29 (ref) | 1.00 | - | 1.00 | - | 1.00 | - |
| 30-39 | 1.21 (1.00, 1.46) | .05 | 1.42 (1.24, 1.63) | <.001 | 0.81 (0.68, 0.96) | .01 |
| 40-49 | 1.34 (1.10, 1.64) | .004 | 1.81 (1.57, 2.07) | <.001 | 1.00 (0.84, 1.20) | .96 |
| 50-64 | 1.47 (1.20, 1.80) | <.001 | 3.04 (2.65, 3.49) | <.001 | 1.15 (0.97, 1.36) | .11 |
| 65-74 | 0.95 (0.75, 1.19) | .64 | 12.76 (10.81, 15.05) | <.001 | 1.04 (0.82, 1.32) | .72 |
| 75+ | 1.55 (1.13, 2.12) | .01 | 25.73 (20.29, 32.67) | <.001 | 0.90 (0.60, 1.33) | .59 |
| **P for trend** |  | .12 |  | <.001 |  | .57 |
| Race/ethnicity |  |  |  |  |  |  |
| Non-Hispanic White (ref) | 1.00 | - | 1.00 | - | 1.00 | - |
| Non-Hispanic Black | 0.92 (0.76, 1.10) | .36 | 1.07 (0.95, 1.20) | .27 | 0.82 (0.42, 0.54) | .02 |
| Non-Hispanic Asian | 1.79 (1.48, 2.18) | <.001 | 1.28 (1.10, 1.50) | .001 | 2.37 (1.41, 1.99) | <.001 |
| Non-Hispanic multiracial | 0.93 (0.74, 1.17) | .55 | 0.98 (0.81, 1.18) | .81 | 0.85 (0.54, 0.79) | .17 |
| Hispanic | 1.06 (0.88, 1.26) | .56 | 1.21 (1.08, 1.36) | .001 | 1.49 (1.07, 1.35) | <.001 |
| Sex |  |  |  |  |  |  |
| Male (ref) | 1.00 | - | 1.00 | - | 1.00 | - |
| Female | 1.69 (1.53, 1.88) | <.001 | 1.30 (1.22, 1.39) | <.001 | 0.81 (0.74, 0.89) | <.001 |
| Education |  |  |  |  |  |  |
| <High school | 0.41 (0.27, 0.63) | <.001 | 0.29 (0.24, 0.36) | <.001 | 0.51 (0.40, 0.64) | <.001 |
| High school | 0.40 (0.34, 0.48) | <.001 | 0.41 (0.37, 0.45) | <.001 | 0.45 (0.40, 0.52) | <.001 |
| Some college | 0.76 (0.69, 0.85) | <.001 | 0.58 (0.54, 0.62) | <.001 | 0.57 (0.51, 0.63) | <.001 |
| ≥College (ref) | 1.00 | - | 1.00 | - | 1.00 | - |
| **P for trend** |  | <.001 |  | <.001 |  | <.001 |
| Household income (2019) |  |  |  |  |  |  |
| <\$25,000 | 0.53 (0.36, 0.77) | .001 | 0.66 (0.54, 0.81) | <.001 | 0.81 (0.60, 1.09) | .17 |
| \$25,000-\$34,999 | 0.63 (0.46, 0.87) | .005 | 0.86 (0.71, 1.04) | .13 | 0.72 (0.53, 0.99) | .04 |
| \$35,000-\$49,999 | 0.68 (0.51, 0.91) | .01 | 0.87 (0.73, 1.04) | .13 | 0.69 (0.52, 0.92) | .01 |
| \$50,000-\$74,999 | 0.78 (0.62, 0.98) | .04 | 0.91 (0.78, 1.07) | .26 | 0.73 (0.56, 0.96) | .03 |
| \$75,000-\$99,999 | 0.79 (0.63, 0.997) | .047 | 0.91 (0.77, 1.08) | .30 | 0.73 (0.56, 0.95) | .02 |
| \$100,000-\$149,999 | 0.96 (0.78, 1.17) | .66 | 1.00 (0.87, 1.16) | .96 | 0.79 (0.61, 1.01) | .06 |
| \$150,000-\$199,999 | 1.06 (0.83, 1.34) | .65 | 0.98 (0.82, 1.18) | .84 | 0.78 (0.58, 1.05) | .11 |
| ≥\$200,000 (ref) | 1.00 | - | 1.00 | - | 1.00 | - |
| **P for trend** |  | .01 |  | .001 |  | .13 |
| Financial hardship |  |  |  |  |  |  |
| Not at all difficult (ref) | 1.00 | - | 1.00 | - | 1.00 | - |
| A little difficult | 0.89 (0.78, 1.00) | .06 | 0.95 (0.87, 1.03) | .21 | 1.01 (0.89, 1.15) | .89 |
| Somewhat difficult | 0.69 (0.59, 0.80) | <.001 | 0.80 (0.72, 0.88) | <.001 | 0.85 (0.73, 0.99) | .03 |
| Very difficult | 0.56 (0.44, 0.70) | <.001 | 0.65 (0.56, 0.75) | <.001 | 0.76 (0.63, 0.90) | .002 |
| **P for trend** |  | <.001 |  | <.001 |  | <.001 |
| Marital status |  |  |  |  |  |  |
| Married (ref) | 1.00 | - | 1.00 | - | 1.00 | - |
| Widowed | 0.81 (0.61, 1.08) | .15 | 0.78 (0.65, 0.94) | .01 | 0.79 (0.59, 1.08) | .14 |
| Divorced/Separated | 0.99 (0.85, 1.16) | .92 | 0.82 (0.74, 0.91) | <.001 | 0.97 (0.84, 1.12) | .70 |
| Never Married | 0.80 (0.68, 0.94) | .01 | 0.82 (0.73, 0.92) | <.001 | 1.12 (0.97, 1.30) | .12 |

Abbreviations: CI, confidence interval; COVID-19, coronavirus disease 2019.
<sup>a</sup> All models included the above predictor variables, and were also adjusted for household size, presence of children in household, and state of residence. All estimates are for adults age 18-85 years. Financial hardship refers to difficulty in paying usual household expenses over the previous week.
<sup>b</sup> Among those not yet vaccinated.
<sup>c</sup> Odds ratio point estimates, 95% confidence intervals, and *P* values were derived from logistic regression models fit using generalized estimating equations that incorporated person weights and robust standard errors. Missing data were handled using 20 multiple imputation data sets.

Furthermore, both education and pre-pandemic income levels exhibited evidence of positive dose-response relationships with vaccine initiation (*P* for linear trend ≤.01 and <.001, respectively). Compared to those with at least a college education, those with less than a high school education were 59-71% less likely to have been vaccinated (*P*<.001). In addition, compared to those with pre-pandemic income levels ≥$200,000, those with income levels <$25,000 were 34-47% less likely to have been vaccinated (*P*<.001). Substantial (vs. no) financial hardship was further linked to a 35-44% lower odds of vaccination (*P*<.001).

Among those not yet vaccinated, qualitatively similar patterns for race/ethnicity, pre-pandemic income, education, and financial hardship during the pandemic were primarily observed for the outcome of a definite intention to receive the vaccine (*Table 1*). However, unlike for vaccine initiation, Black race/ethnicity was associated with a 18% lower likelihood of vaccine intention than White race/ethnicity (*P*=.02).

The most common reasons for vaccine hesitancy were concerns about side effects (Whites—48.0%; Blacks—46.0%; Hispanics—48.6%), and safety (Whites—42.2%; Blacks—49.0%; Hispanics—42.6%) (*Figure 3*). There was evidence of higher levels of concerns about vaccine safety among Blacks vs. Whites. Hispanics, Blacks, and Asians all expressed lower levels of distrust in the government and lower levels of a belief of not needing the vaccine than Whites (*Figure 3*).

**Figure 3.**
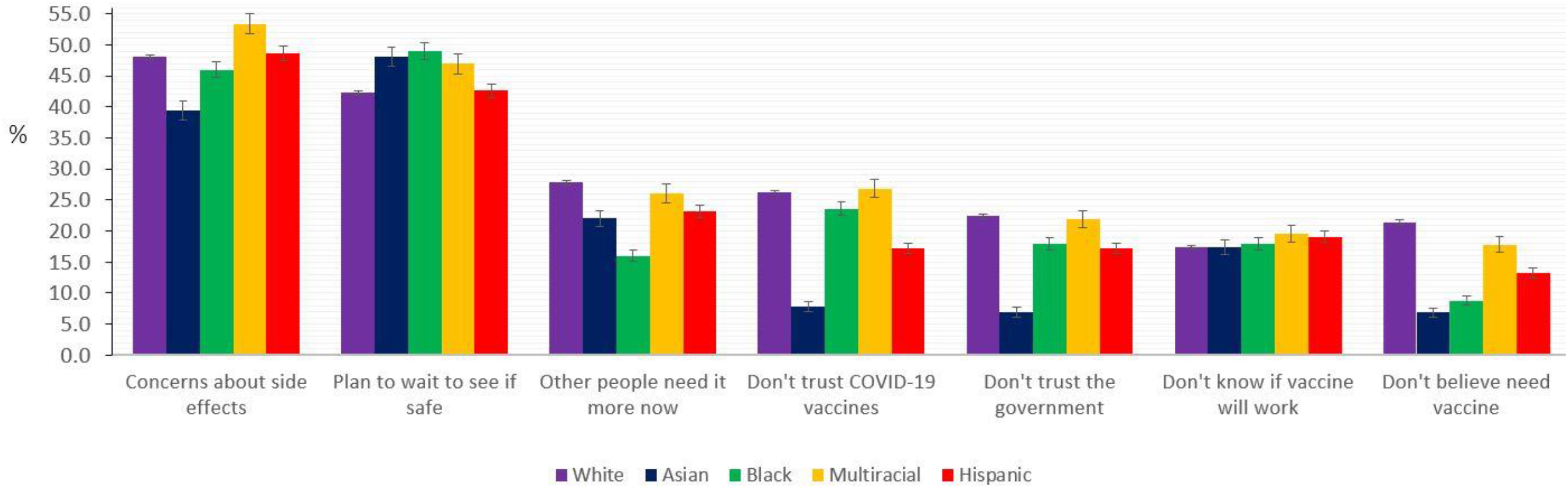
**Reasons for Not Receiving or Planning to Receive COVID-19 Vaccine in Adults Aged 18-85 Years by Race/Ethnicity, U.S. Census Bureau Household Pulse Survey, March 2021.^a^** Abbreviations: COVID-19, coronavirus disease 2019. ^a^ Aggregate data were drawn from the U.S. Census Bureau Household Pulse Survey public-use data tables for surveys administered between March 17, 2021 and March 29, 2021. Error bars indicate 95% confidence intervals for prevalence estimates. All estimates are survey weighted.

## Discussion

In this large, nationally-representative study with relatively complete race/ethnicity and socioeconomic data, we find persistent disparities in vaccine initiation for Blacks, Hispanics, and multiracial persons, and in vaccine intention for Blacks and multiracial persons, compared to Whites and Asians, more than three months after COVID-19 vaccines first became available. To our knowledge, this study is the first to heed recent calls for more complete race and ethnicity data to identify and characterize racial/ethnic disparities in COVID-19 vaccination over time.^1,2^ By the latter half of March 2021, the estimated percentage of Blacks receiving a vaccine was 8 percentage points lower than for Whites. For Hispanics, levels of vaccine intention exceeded those for Whites by late March 2021; however, the percentage with vaccine initiation then was still more than 12 percentage points lower than for Whites. These substantial disparities were present despite the higher representation of Blacks and Hispanics in the vaccination priority groups of healthcare and frontline and other essential workers (Phases 1a, 1b, and 1c) in comparison to the entire U.S. population.^9,10^ For Hispanics, the disparities could possibly reflect a lack of access to vaccines. After adjusting for other factors, having the least education and income and experiencing substantial financial hardship during the pandemic were each independently associated with markedly lower odds of vaccine initiation and vaccine intention.

Controlling for demographic and socioeconomic factors, Black race/ethnicity was linked to a lower vaccine intention but not vaccine initiation than White race/ethnicity i.e., the observed disparities in vaccine initiation between Blacks and Whites did not remain when the distribution of demographic and socioeconomic factors were set to be equal in the two groups. By contrast, in the same models, relatively consistent dose-response relationships were observed for socioeconomic factors and financial hardship. In keeping with causal interpretations of race/ethnicity in multivariable regression models,^11^ these findings would appear to suggest that socioeconomic/economic factors largely account for the observed disparities in vaccine initiation for Blacks vs. Whites.

Study limitations include its cross-sectional and observational design, which cannot rule out bias due to confounding. While sampling weights and raking accounted for non-response and undercoverage, the low survey response rate could have led to selection bias. Last, all measures were based on self-report, and could be subject to social desirability bias for reporting vaccination.^12^

Despite the inherent limitations of this study, its core findings still underscore major inequities in vaccine initiation by race/ethnicity (particularly for Blacks, Hispanics, and multiracial persons), socioeconomic position, as well as financial hardship during the pandemic. In the ensuing months of the vaccine rollout, addressing such persevering racial/ethnic and socioeconomic inequities in vaccination due to lack of access and vaccine hesitancy is critical to mitigate the pandemic’s disproportionately higher risks for infection and adverse health outcomes in Blacks and socioeconomically-disadvantaged groups and to help maximize vaccination coverage nationwide.^1,6^ In addition to the ongoing work of the CDC with jurisdictions to bring vaccines to socially vulnerable communities,^1^ the federal government’s capacity to provide adequate economic relief to reduce levels of financial hardship could play a vital role in improving overall vaccination rates and reducing racial/ethnic and socioeconomic inequities in vaccination, while at the same time attenuating negative impacts on the mental and physical health and social needs of millions of Americans.^13^

## Data Availability

All data are publicly available.

